# Current but Not Former Smoking Is Associated With Higher HbA1c in Adults Without Diabetes

**DOI:** 10.64898/2026.04.10.26350673

**Authors:** Chibuzo C. Manafa, Patrick Onochie Manafa, Nnamdi Okoli, Chukwuzitelu O. Okafor-Udah, Stephen Adilih, Ngozi Ogo, Ndidi-amaka Amy Adilih

## Abstract

**Aim:** We examined associations between smoking and HbA1c among U.S. adults, and whether these associations vary by diabetes status.

**Methods:** We analyzed NHANES data from 2015–2018 for adults aged ≥20 years. Smoking was assessed by self-report and serum cotinine. Survey-weighted multivariable linear regression was used to evaluate the association between smoking and HbA1c in the full population (N=9,214) and in adults without diabetes (N=7,328), adjusting for demographics, blood pressure, waist circumference, lipids, and C-reactive protein.

**Results:** After adjustment for cardiometabolic covariates, there was no significant association between smoking and HbA1c in the full population (former: β=0.029%, p=0.30; current: β=0.053%, p=0.13). Among adults without diabetes, former smoking was not associated with HbA1c, whereas current smoking remained significantly associated (former: β=−0.001%, p=0.923; current: β=0.067%, p<0.001). These findings were similar when cotinine was used as the exposure measure, with active smoking (≥3.0 ng/mL) associated with higher HbA1c among non-diabetic adults (p<0.001), but not in the full population.

**Conclusions:** Among adults without diabetes, current but not former smoking was associated with higher HbA1c. The absence of an association in former smokers suggests that this effect may attenuate following cessation. These findings support early cessation interventions and may inform cessation counseling and diabetes screening.

## INTRODUCTION

Cigarette smoking is a well-known risk factor for type 2 diabetes mellitus, and meta-analyses have noted an increased risk of about 30–50% among current smokers when compared to never smokers [1,2]. Previous studies have consistently reported higher HbA1c levels among smokers [3,4,5]. However, the biological pathways underlying this association and its clinical implications remain incompletely understood.

Glycated hemoglobin (HbA1c) is a measure of average glycemia over 2–3 months, corresponding to the lifespan of red blood cells. Factors other than glucose concentration, such as red blood cell turnover, hemoglobin characteristics, systemic inflammation, and adiposity, may affect HbA1c levels [6,7]. Since diabetes screening measures rely on strict HbA1c thresholds of 5.7% for prediabetes and 6.5% for diabetes [8], a modest increase in HbA1c could influence risk classification; therefore, clarifying the mechanisms underlying the association between smoking and HbA1c is of potential clinical and epidemiological relevance.

The association between smoking and HbA1c may differ based on an individual’s diabetes status, and comparing current with former smokers can help differentiate reversible from more long-term effects. Similar HbA1c levels in former and never smokers would support the reversibility of the effects of smoking, suggesting a more direct effect of smoking on the red blood cells and possibly on insulin resistance [6]. However, persistently elevated levels following smoking cessation could suggest longer-lasting metabolic effects targeting systemic inflammation and adiposity as proposed by Chiolero et al [7].

In their study of individuals with established diabetes, Kar et al. reported that glycemic recovery following cessation may be incomplete and may require several years of sustained abstinence [9]. However, it is still unclear whether similar patterns of association or reversibility are present in adults without diabetes.

Previous studies have mostly relied on self-reported smoking status, but this can be subject to misclassification and social desirability bias [10]. Serum cotinine, the primary metabolite of nicotine with a half-life of 16–18 hours, serves as an objective biomarker of recent tobacco exposure and can be an alternative measure [11]. Evaluating both self-reported smoking status and cotinine concentrations within the same analytical framework may help distinguish between exposure misclassification and true biological associations.

Findings by Clair et al. showed that cotinine was associated with higher HbA1c levels among non-diabetic adults [3]. Given the declining smoking prevalence in recent years [12], the present analysis builds on this foundational work by examining recent National Health and Nutrition Examination Survey (NHANES) cycles.

Using NHANES 2015–2018 data, we examined associations of self-reported smoking status and serum cotinine with HbA1c in U.S. adults. We hypothesized that smoking would be associated with higher HbA1c, that this association would attenuate after adjustment for cardiometabolic covariates, and that associations derived from self-reported smoking and serum cotinine would be concordant. We further hypothesized that, among adults without diabetes, former smokers would demonstrate HbA1c levels comparable to never smokers.

## METHODS

### Study Design and Data Source

We conducted a cross-sectional analysis of data from the National Health and Nutrition Examination Survey (NHANES). This is a nationally representative survey of the noninstitutionalized U.S. population. We pooled two continuous cycles (2015–2016 and 2017–2018) to increase statistical precision.

NHANES protocols were approved by the National Center for Health Statistics Research Ethics Review Board. Every participant also provided informed consent before their data were collected. We did not require any further ethics approval, as the data are publicly available and do not contain any identifiable information.

### Study Population

We restricted our participants to adults aged ≥20 years. The initial sample across the 4 years included 19,225 participants, and we restricted analyses to individuals with complete data on HbA1c, smoking status, waist circumference, blood pressure, lipids, and high-sensitivity C-reactive protein (hsCRP). We conducted a further subgroup analysis among participants without diabetes (Supplementary Figure S1).

We had 9,214 participants for self-reported smoking analyses, 9,221 for cotinine-based analyses, and 7,328 for the non-diabetic subgroup analyses.

### Exposure Assessment

#### Self-Reported Smoking

Smoking status was determined using standardized NHANES questionnaire items (SMQ020 and SMQ040). Participants who had smoked <100 cigarettes in their lifetime were classed as never smokers, and they were used as the reference group. We grouped those participants who had smoked ≥100 cigarettes in their lifetime but are not currently smoking as former smokers, while those who had smoked ≥100 cigarettes in their lifetime and still continue to smoke were classed as current smokers.

#### Serum Cotinine

Serum cotinine concentrations were quantified using isotope-dilution high-performance liquid chromatography coupled with tandem mass spectrometry. Cotinine is a major metabolite of nicotine and is used as a biomarker to check for active smoking. The detection limit was 0.011 ng/mL.

For the main analyses, cotinine levels were categorized into the following cut points: <0.05 ng/mL, 0.05–<3.0 ng/mL, and ≥3.0 ng/mL. We noted that about a third of the cotinine values were at or near the assay detection limit. As a result of this clustering, we analyzed cotinine via multiple methods for sensitivity checks. We analyzed cotinine in quintiles to evaluate whether the association was nonlinear, assessed cotinine as a log-transformed continuous variable, and also performed a winsorized analysis, capping it at the 99th percentile (Supplementary Tables S3–S4).

Concordance between self-reported smoking and cotinine categories was assessed using survey-weighted cross-tabulation.

### Outcome Assessment

Our primary outcome, HbA1c, was analyzed as a continuous variable. HbA1c was measured using high-performance liquid chromatography from whole blood.

#### Covariates

All models included age, sex, and race/ethnicity (RIDRETH3) as baseline, and then cardiometabolic variables were added based on prespecified models. The cardiometabolic variables we used are systolic blood pressure, measured as the mean of readings 2–4; waist circumference; non-HDL cholesterol (total cholesterol minus HDL cholesterol); and high-sensitivity C-reactive protein (hsCRP). hsCRP showed a right-skewed distribution, so we modeled it as log(1 + hsCRP).

The main model used waist circumference and not BMI. This is because waist circumference has been shown to correlate better with cardiovascular risk [13].

#### Diabetes Definition and Subgroup Analysis

Diabetes was defined based on self-reported data or HbA1c. Participants were considered to have diabetes if they had been told by a health professional that they had diabetes or if the HbA1c was ≥6.5%.

For adults without diabetes, we repeated the primary model, restricting it to participants who had not been told by a health professional that they had diabetes and those with HbA1c <6.5% (N=7,328).

### Statistical Analysis

All analyses accounted for NHANES’ complex survey design using examination weights (WTMEC2YR), primary sampling units (SDMVPSU), and strata (SDMVSTRA). All data were analyzed using R version 4.5.2. Following the recommendations by the National Center for Health Statistics, four-year weights were constructed by dividing the 2-year examination weights by 2. Variance estimation used Taylor series linearization with adjustment for strata containing a single primary sampling unit.

We built different survey-weighted linear regression models to assess for associations between smoking exposure and HbA1c. In the first model, we adjusted for age, sex, and race/ethnicity, which served as our base. For the second model, we added systolic blood pressure. For the third and fourth models, we examined BMI and waist circumference separately, each building on Model 2. The fifth model included both BMI and waist circumference. Model 6 was prespecified as the primary model, and it extended the waist-circumference-based model by including non-HDL cholesterol and log-transformed hsCRP. Model 7 additionally adjusted for education and the poverty–income ratio, but served as a sensitivity model due to many more missing values.

For models with BMI included, the sample size was slightly smaller due to missing data (N=9,198). Model 7 was estimated using complete-case analysis (N=8,151).

We conducted a formal test for interaction between smoking and diabetes status by including a smoking × diabetes term in our fully adjusted model.

Overall, using survey-weighted proportions, we found that missingness was low (2.3–5.8%) and there was very little variation across smoking categories (<3%), supporting the use of complete-case analysis (Supplementary Table S1).

Statistical significance was defined as a two-sided p<0.05.

### Transparency and Reproducibility

The R code and the session information are all available upon request.

## Results

### Study Population Characteristics

From an initial sample of 19,225 NHANES participants, we excluded participants aged <20 years. Among the remaining participants, we further excluded those with missing data (Supplementary Figure S1). A total of 9,214 adults were included in the self-reported smoking analyses, 9,221 in the cotinine analyses, and 7,328 adults without diabetes in the subgroup analyses. Missingness across variables was low (2.3–5.8%) and similar across smoking groups (Supplementary Table S1). All analyses incorporated NHANES survey weights.

Weighted prevalence of smoking categories was 56.5% never, 25.5% former, and 17.9% current smokers (Supplementary Figure S1). Current smokers were younger (mean age 44.3 years) than never smokers (46.8 years) and former smokers (53.7 years) and had lower educational attainment. Mean HbA1c values were 5.62% (SE 0.02), 5.79% (SE 0.03), and 5.65% (SE 0.03) among never, former, and current smokers, respectively. Former smokers had greater waist circumference and systolic blood pressure than the other groups. Former and current smokers were more likely to be male compared with never smokers (Table 1).

**Table 1.** Baseline Characteristics by Self-Reported Smoking Status (N=9,214)

| **Characteristic** | **Never** | **Former** | **Current** |
| --- | --- | --- | --- |
| **Continuous Variables – Weighted Mean (SE)** |  |  |  |
| Age, years | 46.80 (0.43) | 53.67 (0.68) | 44.34 (0.67) |
| HbA1c, % | 5.62 (0.02) | 5.79 (0.03) | 5.65 (0.03) |
| BMI, kg/m<sup>2</sup> | 29.41 (0.21) | 30.50 (0.24) | 28.84 (0.25) |
| Waist circumference, cm | 99.22 (0.53) | 105.09 (0.60) | 99.73 (0.69) |
| Systolic BP, mmHg | 122.30 (0.36) | 126.21 (0.69) | 122.52 (0.76) |
| Diastolic BP, mmHg | 71.36 (0.39) | 71.24 (0.52) | 71.59 (0.64) |
| Non-HDL cholesterol, mg/dL | 134.26 (1.11) | 139.34 (1.79) | 139.29 (1.79) |
| High sensitivity C-reactive protein, mg/L | 3.55 (0.13) | 3.97 (0.21) | 4.31 (0.24) |
| **Categorical Variables – Weighted %** |  |  |  |
| **Sex** |  |  |  |
| Male | 41.4% | 60.9% | 54.4% |
| Female | 58.6% | 39.1% | 45.6% |
| **Race/Ethnicity** |  |  |  |
| Mexican American | 10.3% | 7.0% | 7.3% |
| Other Hispanic | 7.4% | 5.4% | 5.0% |
| Non-Hispanic White | 60.4% | 73.3% | 64.1% |
| Non-Hispanic Black | 11.3% | 6.9% | 13.6% |
| Non-Hispanic Asian | 7.7% | 2.7% | 2.5% |
| Other / Multiracial | 2.9% | 4.7% | 7.5% |
| **Education** |  |  |  |
| Less than high school | 10.4% | 12.0% | 18.2% |
| High school / GED | 20.1% | 26.2% | 34.8% |
| Some college | 30.0% | 34.8% | 34.6% |
| College graduate or higher | 39.4% | 27.1% | 12.5% |

**Abbreviations:** BMI, body mass index; HbA1c, hemoglobin A1c; HDL, high-density lipoprotein; Non-HDL, non-HDL cholesterol; BP, blood pressure.
Values are weighted means (SE) or percentages. Percentages sum to 100% within smoking category.

### Self-Reported Smoking Status and HbA1c

In Model 1, adjusted for only demographics, both former and current smoking were associated with higher HbA1c compared with never smoking (former: β=0.078%, 95% CI 0.016 to 0.140; current: β=0.077%, 95% CI 0.018 to 0.135). When adjusted for systolic blood pressure, there was minimal change in effect estimates (Model 2).

Addition of BMI reduced the association for former smoking but not for current smoking. However, after the inclusion of waist circumference, non-HDL cholesterol, and log(1+hsCRP) as shown in model 6, effect estimates decreased and were no longer statistically significant (former: β=0.029%, 95% CI −0.028 to 0.085; current: β=0.053%, 95% CI −0.016 to 0.121).

Model 7 was added as a sensitivity model as it had fewer participants due to missing values (n=8151). It adjusted for education and poverty-income ratio but did not materially change these findings (Table 2). Age, waist circumference, non-HDL cholesterol, and log(1+hsCRP) remained independently associated with HbA1c (Supplementary Table S2).

**Table 2.** Sequential Models - Association Between Self-Reported Smoking Status and HbA1c (N=9,214)

| **Model** | **Covariates** | **Former vs Never**<br>**$\beta$ (95% CI)** | **p** | **Current vs Never**<br>**$\beta$ (95% CI)** | **p** |
| --- | --- | --- | --- | --- | --- |
| M1 | Age + Sex + Race/Ethnicity | 0.078 (0.016, 0.140) | 0.016 | 0.077 (0.018, 0.135) | 0.012 |
| M2 | Model 1 + SBP | 0.075 (0.011, 0.138) | 0.024 | 0.073 (0.015, 0.130) | 0.016 |
| M3 | Model 2 + BMI | 0.048 (-0.013, 0.109) | 0.113 | 0.093 (0.030, 0.155) | 0.006 |
| M4 | Model 2 + Waist Circumference | 0.035 (-0.024, 0.094) | 0.233 | 0.078 (0.016, 0.140) | 0.017 |
| M5 | Model 2 + BMI + Waist Circumference | 0.033 (-0.026, 0.093) | 0.257 | 0.077 (0.011, 0.142) | 0.025 |
| **M6** | **Model 4 + Non-HDL + log(hsCRP)** | **0.029 (-0.028, 0.085)** | **0.301** | **0.053 (-0.016, 0.121)** | **0.125** |
| M7 | Model 6 + Education + PIR | 0.028 (-0.037, 0.093) | 0.363 | 0.033 (-0.049, 0.116) | 0.398 |

**Abbreviations:** BMI, body mass index; CI, confidence interval; HbA1c, hemoglobin
A1c; HDL, high-density lipoprotein; hsCRP, high-sensitivity C-reactive protein;
Non-HDL, non-HDL cholesterol; PIR, poverty-income ratio; SBP, systolic blood pressure.
Note: $\beta$ coefficients represent adjusted differences in HbA1c (%) relative to never smokers. Model 6 is the primary fully adjusted model. Model 7 includes education and PIR; N=8,151 due to missing socioeconomic data (complete-case analysis). Never smokers are the reference group.

### Associations by Exposure Measure and Diabetes Status

After full adjustment in the overall population, there was no association between smoking and HbA1c, and this was true for both self-reported smoking and cotinine-defined exposure (Table 3). However, among participants without diabetes (n=7,328), current smoking was associated with higher HbA1c (β=0.067%, 95% CI 0.039 to 0.094, p<0.001), whereas former smoking was not (β=−0.001%, 95% CI −0.027 to 0.025, p=0.923). Associations did not show a significant difference by diabetes status (former p=0.375; current p=0.808). Also, higher HbA1c was observed among participants with cotinine levels consistent with active smoking (≥3.0 ng/mL), but not among those showing environmental exposure levels (β=0.064%, 95% CI 0.036 to 0.093, p<0.001) (Table 3, Figure 1).

**Table 3.** Association Between Smoking and HbA1c by Exposure Measure and Diabetes Status.

| Exposure | Full Population |  | Non-Diabetic Adults |  |
| --- | --- | --- | --- | --- |
| | $\beta$ (95% CI) | P value | $\beta$ (95% CI) | P value |
| **Self-Reported Smoking Status** |  |  |  |  |
| Former vs Never | 0.029 (−0.028, 0.085) | 0.301 | −0.001 (−0.027, 0.025) | 0.923 |
| Current vs Never | 0.053 (−0.016, 0.121) | 0.125 | 0.067 (0.039, 0.094) | <0.001 |
| **Serum Cotinine Categories** |  |  |  |  |
| 0.05–<3.0 ng/mL (ETS) vs <0.05 | 0.004 (−0.053, 0.062) | 0.880 | 0.021 (−0.007, 0.048) | 0.130 |
| ≥3.0 ng/mL (Active) vs <0.05 | 0.036 (−0.037, 0.108) | 0.313 | 0.064 (0.036, 0.093) | <0.001 |

**Abbreviations:** HbA1c, glycated hemoglobin; ETS, environmental tobacco smoke; CI, confidence interval.
$\beta$ coefficients represent percentage-point differences in HbA1c from fully adjusted models.
Sample sizes: N=9,214 (self-reported smoking), N=9,221 (serum cotinine), N=7,328 (non-diabetic subsample)

**Figure 1.**
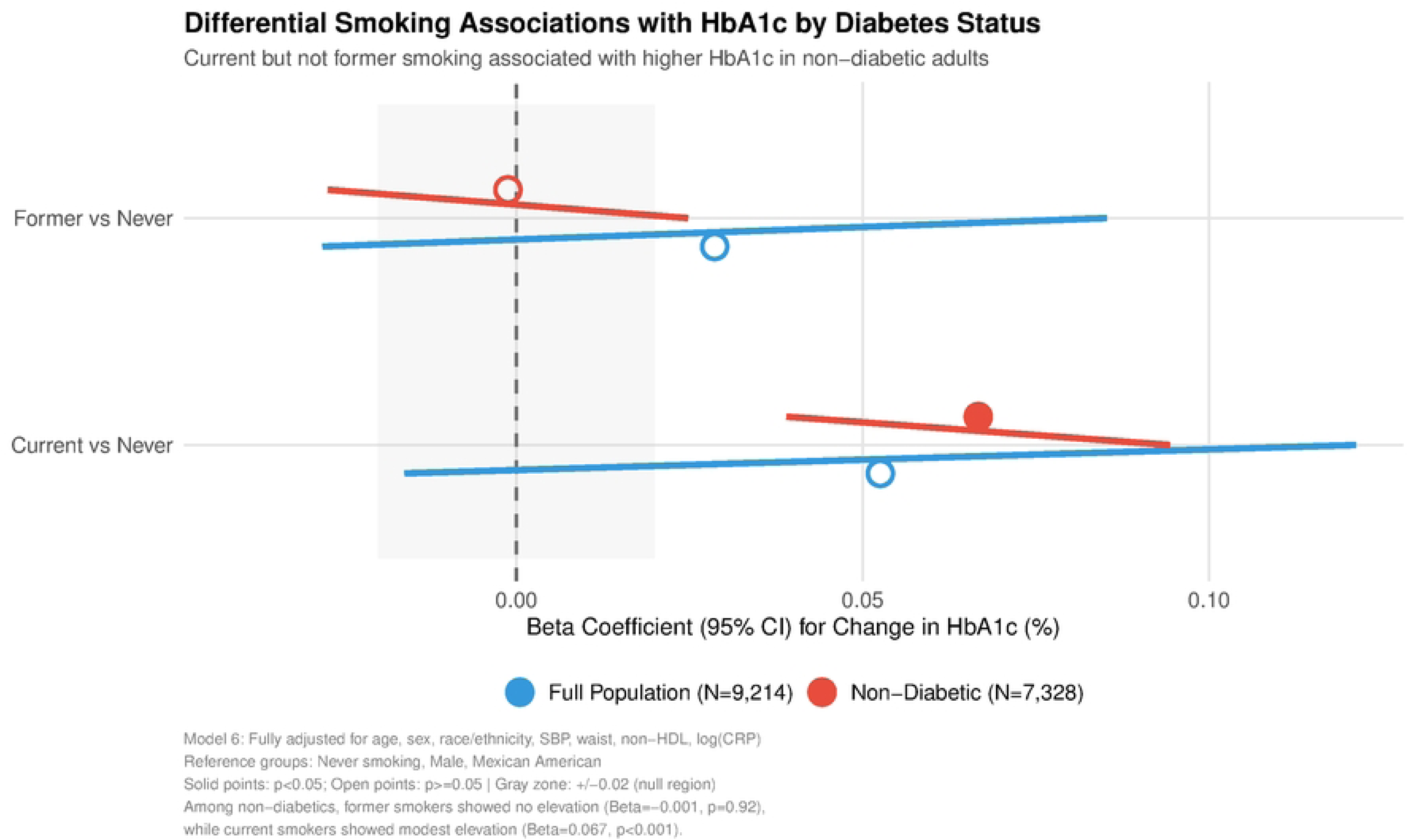
Differential Smoking Associations with HbA1c by Diabetes Status. Comparison of smoking associations with HbA1c in full population (N=9,214) versus participants without diabetes (N=7,328). Effect estimates from fully adjusted Model 6. Diabetes defined as self-reported diagnosis or HbA1c ≥6.5%. Solid points: p<0.05; open points: p≥0.05. Gray zone: ±0.02% HbA1c (approximate null region).

Approximately one-third of cotinine values (34.2%) were at the detection limit (0.011 ng/mL). Regardless of how cotinine was modeled—whether by categories, quintiles, or as a continuous log-transformed variable—no monotonic dose–response relationship with HbA1c was observed (p for trend=0.217). Estimates were similar after winsorization at the 99th percentile (Supplementary Tables S3–S4).

### Concordance Between Self-Reported Smoking and Serum Cotinine

There was high overall concordance between self-reported smoking and cotinine categories. Cotinine levels consistent with active smoking were observed in 6.1% of never smokers and 18.1% of former smokers (Supplementary Table S5).

## Discussion

In this study, we analyzed NHANES data from 2015–2018. This represents an ethnically diverse cohort of U.S. adults. We found that the association between smoking and HbA1c was largely attenuated after comprehensive adjustment for central adiposity, inflammatory markers, and lipid measures in the full population. Further subgroup analyses showed that among participants without diabetes, there was a modest rise in HbA1c for current smokers, while former smokers had similar values to never smokers.

In a prior NHANES analysis, Clair et al. reported a positive association between smoking and HbA1c after adjustment for multiple covariates [3]. Similar findings were observed in the EPIC-Norfolk study and in analyses of the Korea National Health and Nutrition Examination Survey [4,5]. However, these studies did not adjust simultaneously for adiposity, inflammatory markers, and lipid profiles. To contextualise this difference, we reconstructed adjustment strategies similar to those in previous studies. Our Model 4, which included demographics, blood pressure, and waist circumference, mirrored previous reports and yielded similar findings. Once inflammatory indices and lipid measures were added, the association attenuated further and was no longer statistically significant. We observed a decrease of approximately 63% in the coefficient for former smokers and about one-third for current smokers from the base model to the fully adjusted model (Model 6) (Supplementary Figure S2). This pattern suggests that the associations observed in minimally adjusted models may reflect clustering of metabolic and inflammatory pathways rather than a direct, independent effect of smoking on glycemia.

Adjustment for waist circumference attenuated the association more than BMI, and the model including both measures yielded estimates similar to the waist circumference–adjusted model. This suggests that central adiposity, as measured by waist circumference, is the primary adiposity-related pathway. Interestingly, for current smokers, adjusting for BMI alone increased the coefficient, reflecting the lower body weight commonly observed in active smokers, as noted by Chiolero et al [7].

In the subgroup analyses, after full adjustment for blood pressure, adiposity, inflammatory markers, and lipid measures, HbA1c levels among former smokers were similar to those of never smokers. This pattern suggests that the effect of smoking on HbA1c may diminish following cessation, although the cross-sectional design does not allow us to determine timing or direction. This pattern was observed despite former smokers having higher adiposity than never smokers (BMI 30.50 vs. 29.41 kg/m²; waist circumference 105.09 vs. 99.22 cm). These findings were consistent when cotinine concentrations were used as the exposure metric (β=0.064%, p<0.001).

The smoking-diabetes interaction test did not reach statistical significance. Nonetheless, subgroup analyses suggested results compatible with known biological mechanisms. As such, this merits replication in larger sample studies with adequate power.

We observed a modest discordance between self-reported smoking and cotinine categories. This could be as a result of relapse, intermittent smoking, or social desirability bias [10]. Nevertheless, findings were consistent across both exposure definitions. Only active smoking (≥3.0 ng/mL) was associated with higher HbA1c, whereas environmental tobacco smoke exposure was not. The quintile and log-adjusted cotinine sensitivity tests did not indicate a dose-dependent relationship either. These findings together support a threshold rather than a dose-dependent relationship.

Our finding contrasts with longitudinal evidence from populations with established diabetes. Kar et al., in a meta-analysis of 98,978 individuals with diabetes, reported HbA1c levels that were 0.61% higher in current smokers than in non-smokers, noting that it may take up to 10 years after quitting for HbA1c levels to converge [9]. Similarly, Lycett et al. described an initial worsening of HbA1c among smokers with diabetes during the first year after cessation [14]. In contrast, we did not observe residual elevation among non-diabetic former smokers, suggesting that smoking-associated differences in HbA1c may vary by diabetes status, although this requires confirmation in longitudinal studies.

Several mechanisms may contribute to these observations. Smoking induces systemic inflammation and oxidative stress, impairing insulin signaling and promoting insulin resistance [15,16]. Our study found that the greatest attenuation occurred after adjusting for central adiposity. Waist circumference alone was associated with approximately 58% of the observed attenuation in the former smoking group. Non-HDL cholesterol and hsCRP led to further attenuation, although to a lesser degree than adiposity (Supplementary Figure S2). HsCRP also demonstrated the strongest independent association with HbA1c among all covariates in the fully adjusted model (β=0.141, p<0.001), suggesting that inflammation is strongly associated with glycemia, despite its effect being secondary to visceral adiposity. Of note, hsCRP levels in former smokers remained modestly higher than those of never smokers, raising the possibility that improvements in glycemia may occur despite residual low-grade inflammation. This also mirrors several prior reports suggesting that inflammatory response resolution may lag behind after smoking cessation, even when other metabolic parameters have stabilized [17–19].

Functional β-cells in early phases of metabolic dysfunction may trigger insulin release to counter some of the effects of smoking on glycemia. This may explain the mild hyperglycemia observed despite hyperinsulinemia in early insulin resistance [20]. Bergman et al., in their experimental work, demonstrated that smoking-induced insulin resistance operates primarily by impairing downstream insulin signaling, which in turn reverses partially within weeks of abstaining from smoking [6]. Given that HbA1c measures average glycemia over 2–3 months, even partial recovery of insulin sensitivity over that period may be enough to reduce HbA1c levels despite post-cessation weight gain, explaining the absence of HbA1c elevation observed in non-diabetic former smokers.

This study has several strengths. The use of nationally representative NHANES data with complex survey weighting enhances generalizability to the U.S. adult population. Our demonstration that objective serum cotinine measurement validates self-reported smoking classification means that both can be used as exposure methods for analysis of the effects of smoking. Our sequential modeling enabled us to measure the attributes of each covariate, providing a more comprehensive analysis than previously reported. Stratification by diabetes status also revealed heterogeneity not present in the full population, and the validation of both self-reported and cotinine measures strengthens the robustness of these associations.

From a public health perspective, these findings reinforce the importance of early smoking cessation interventions. The absence of HbA1c elevation among non-diabetic former smokers, despite their higher adiposity compared to never smokers, suggests that metabolic recovery is achievable following cessation. This may provide additional motivation for smoking cessation counseling, particularly in individuals with other cardiometabolic risk factors. The use of serum cotinine to validate self-reported smoking status strengthens the evidence base and confirms that environmental tobacco smoke exposure, unlike active smoking, is not associated with measurable HbA1c changes in this population.

This study has several limitations. First, the cross-sectional design precludes causal inference. In addition, the NHANES design limited our ability to measure smoking cessation timelines or smoking intensity. Our test for interaction did not reach statistical significance; therefore, the results should be interpreted cautiously. Residual confounding from diet, physical activity, and alcohol use, which were not incorporated into our models, is also possible.

From a clinical perspective, while the increase in HbA1c among current smokers without diabetes may appear modest, it may nonetheless influence patient classifications. This is especially important around diagnostic thresholds. Approximately 13.7% of all U.S. adults are active smokers [12], meaning that small elevations in HbA1c could classify some individuals into the prediabetes range, especially if smoking status is not accounted for in risk-stratification models. Finally, the absence of residual HbA1c elevation among former smokers further supports emphasizing early smoking cessation, particularly prior to the onset of established diabetes.

## Data Availability

Data are publicly available from the National Health and Nutrition Examination Survey (NHANES) conducted by the Centers for Disease Control and Prevention (CDC). The specific datasets analyzed in this study are from the NHANES 2015-2016 and 2017-2018 continuous cycles and can be accessed at https://www.cdc.gov/nchs/nhanes/index.htm. No special permissions are required to access these data.

## Personal Thanks

None.

## Funding and Assistance

None.

## Author Contributions and Guarantor Statement

**C.C.M.** conceived the study, researched data, contributed to the discussion, and wrote the first draft of the manuscript. **P.O.M.** contributed to the discussion and reviewed and revised the manuscript for important intellectual content. **Nnamdi O. (N.O.)**, **C.O.O** and **S.A.** researched data and reviewed and edited the manuscript. **Ngozi Ogo (N.Og.)** and **N.A.A.** contributed to the discussion and reviewed and edited the manuscript. All authors approved the final version of the manuscript.

**C.C.M.** is the guarantor of this work and, as such, had full access to all the data in the study and takes responsibility for the integrity of the data and the accuracy of the data analysis.

## Prior Presentation

None.

